# mouth-to-mouth ventilation through cardiopulmonary resuscitation, is there any other way?

**DOI:** 10.1101/2021.03.04.21252899

**Authors:** Aini Maimaitiming, Xiaohai Wang

## Abstract

**objective:** to provide and explore possibility of new idea that perform mouth-to-mouth ventilation through cardiopulmonary resuscitation.

**Methods:** stage one is establishing the ventilation technique using cola bottle, stage two is measuring the tidal volume when different sized cola bottles were used.

**Result:** the smallest sized cola bottle(500ml) can also make obvious thorax rise in manikin CPR model. The tidal volume is 174.5±9.1ml, 220±7.6ml and 447±15.9ml respectively for 500ml, 600ml and 1.25L cola bottles when using single hand performance. There are statistical differences (0.001) in tidal volume of different sized cola bottle by using one hand performance and two hands.

**Conclusion:** larger sized cola bottles(600ml,1.25L) could be used as substitute ventilation technique for mouth-to-mouth ventilation in special circumnutates.

## Introduction

Mouth-to-mouth ventilation (MMV) has been used in prehospital cardiopulmonary resuscitation (CPR) for decades. Studies have showed that bystander’s willingness toward preforming MMV to stranger is different from each other. Dobbie etc’s^1^ survey showed that 35% respondent’s primary concern about performing CPR is MMV related concerns, despite over half (52%) of them had been trained in CPR courses. One of the main reason holds back trained adult preforming MMV on stranger is fears of contracting infection disease^2, 3^. Another disadvantage of MMV is oxygen content in expired air around 16%-17% much lower than oxygen content in the air (21%), furthermore expired air in MMV also content 4%of carbon dioxide which can easily lead to hypercapnia^4^. In addition to MMV there is some ventilation technique like bag-valve-mask, mouth-to-mask ventilation for use before more permanent (endotracheal intubation) ventilation technique has performed^5^. However these temporal ventilation technique isn’t wildly available for public in real-time out of hospital CPR. The cola bottles, can be found anywhere, have a completely different usage in CPR. Since cola bottles have strong elasticity, it can be sued as simple breathing balloon in the special circumstances. Our aim in this study is to provide new prospect of ventilation technique by using cola bottle for public in CPR and explore its possibility.

## Material and method

Three different sized cola bottles (500ml,680ml,1.25L) were used in this study. In first stage we have performed ventilation technique on manikin CPR model. The cola bottle has put into mouth of manikin model then one hand grab around bottle and hold nose to prevent air leakage while other hand squeezes the middle of the bottle (Fig.1).

**Fig. 1.**
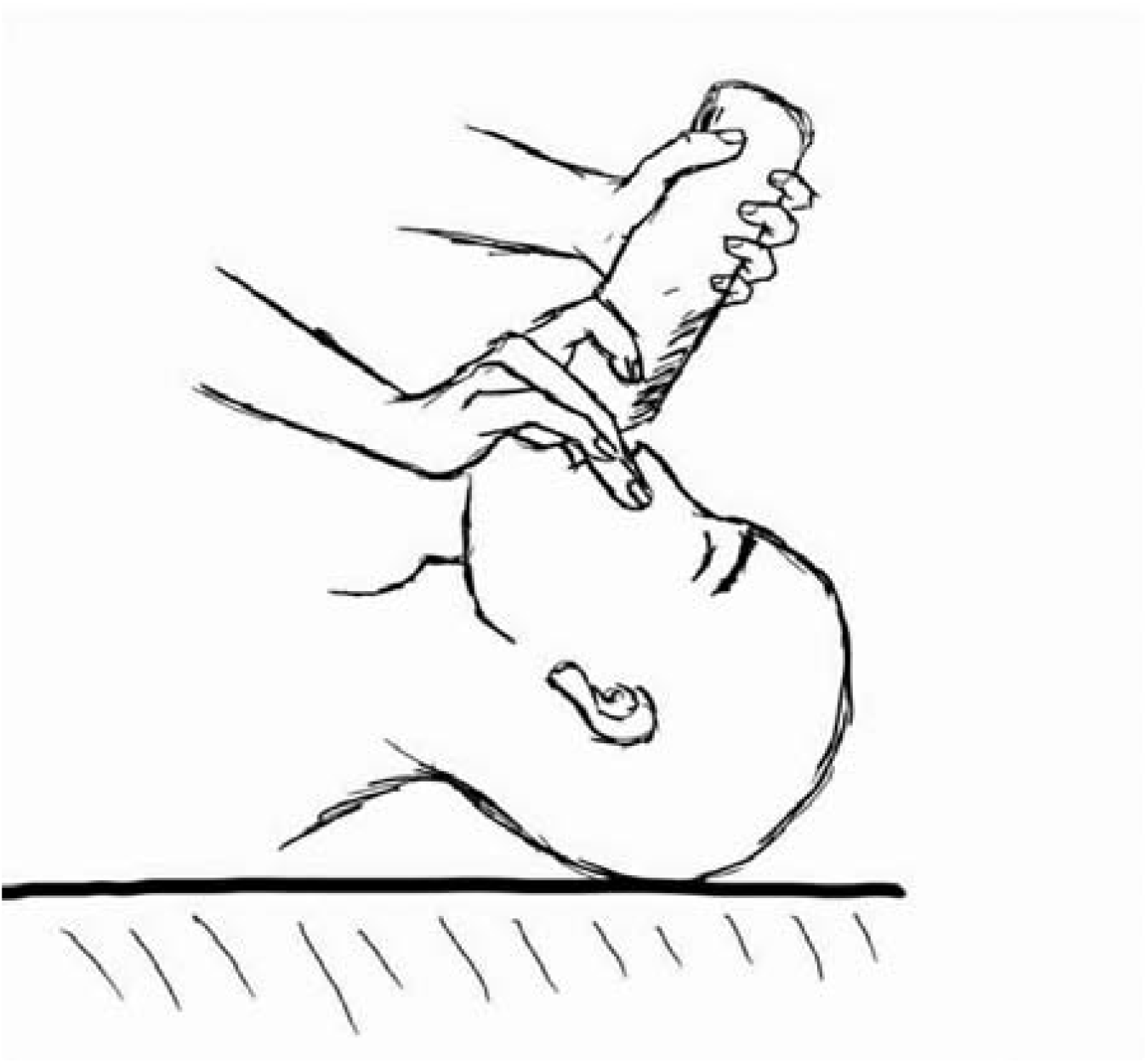
the model of ventilation using cola bottle.

Second stage we connect cola bottles to anesthesia machine (Drager, FabliusGS) to measure tidal volume. The cola bottle installed with airbag connecter and breathing circuit connected to test lung. During operation all gas flowmeter turned off and APL valve adjusted to 30mmH_2_O. Three coworker who has not involved in authorship voluntarily participated in the study. Every participant squeezes middle of the bottle, with full recoil, with one hand and two hands respectively for ten time and tidal volume have recorded.

## Result

Chest arousal have been observed even when the smallest bottle(500ml) is used. The thorax fluctuation is more evident while using the larger cola bottle. There is no statistical difference in there participance’s tidal volume (not given respectively). The tidal volume is 174.5±9.1ml and 218.5± 11.4 for 500ml cola bottle using single hand an both hands respectively, 220±7.6ml, 259.1±21.2ml for 650ml and 447±15.9ml, 477±15.8ml for 1.25L bottle(table.1). There is statistical significance (0.000) in tidal volume which performed single hand and two hands in three bottles. There is no statistical difference (0.087) in tidal volume of 500ml and 650ml cola bottle when technique performed by two hands.

**Table 1.** tidal volume of ventilation when different sized cola bottle and different method is used

| Bottle size | Single hand | Two hands | *P* (between groups) |
| --- | --- | --- | --- |
| 500ml | 174.5±9.1a | 218.5±11.4a | 0.000 |
| 680ml | 220±7.6b | 259.1±21.2ab | 0.001 |
| 1.25L | 447±15.9c | 477±15.8c | 0.000 |
| *P* (between bottles) | 0.000 | 0.000 |  |

All values are given in mean ± standard error of the mean

## DISCUSSION

Is tidal volume which performed by cola bottle enough to maintain the oxygen level and to eliminate carbon dioxide? AHA has suggested that tidal volume should be provided to cause visible chest rise^6^. In our study chest rise has observed even when smallest sized cola bottle(500ml) is used. there have been controversial ideas about optimal ventilation volume during CPR^7-9^. Stallinger A etc’s study^10^ shows that compared to 500ml and 1000ml MTM gas there are higher oxygen saturation and arterial oxygen partial pressure values in using 500ml room air for ventilation and arterial carbon dioxide partial pressure is lower when room air used for ventilation. Furthermore, the study performed on healthy volunteer with normal blood pressure, which means that tidal volume which cardiac arrest victim needed may lower then studied tidal volume because of relatively insufficient blood follow in body.

Experiment can’t be carried in real time out of hospital CPR, therefor there is no standard tidal volume for prehospital resuscitation yet. Since the reason of CPR is to maintain circulation in body to transfer oxygen for the main organs, it is beneficial to ventilate as much as possible within normal limit. Bola bottles(500ml) could not contribute safe tidal volume in pre hospital rescue, still it is batter then no ventilation during prolonged CPR. The public concerns about preforming MTM ventilation can be avoided by using cola bottles in ventilation. Also, it may increase the quality of prehospital ventilation by using larger cola bottles.

## Conclusion

Using cola bottles as simple breathing balloon may increase bystander’s willingness performing ventilation in prehospital CPR. Tidal volume may be sufficient when largest (1.25L) cola bottle is used. However, the positive effects and feasibility of the technique need further examination in real time CPR.

## Data Availability

not applicable

## Acknowledgements

Conception and design of the study, acquisition and interpretation of data, revising the article, final approval of the version to be published: Xiaohai Wang. Acquisition of data, analysis and interpretation of data, drafting the article, final approval of the version to be published: Aini Maimaitiming

## Availability of data and materials

The study data are available on request to the corresponding author

## Informed consent and Ethical approval

The present study did not include the patients or patient’s data.

